# Ensemble Deep Learning for Histopathological Breast Cancer Detection

**DOI:** 10.1101/2025.08.12.25333539

**Authors:** Alireza Rahi

**Affiliations:** Independent Researcher Tehran, Iran

**Keywords:** Breast cancer, Histopathological image classification, BreaKHis dataset, Ensemble deep learning, ResNet50, DenseNet121, Stacking, XGBoost, Voting classifier, Fine-tuning

## Abstract

Breast cancer remains one of the leading causes of mortality among women worldwide, and early and accurate diagnosis is essential for effective treatment. In this study, we propose an ensemble deep learning approach for classifying histopathological images of breast cancer using the BreaKHis dataset. Two state-of-the-art convolutional neural network architectures, ResNet50 and DenseNet121, were fine-tuned and combined through multiple ensemble strategies, including stacking with logistic regression, XGBoost, and hard/soft voting. The models were trained on an 80/20 train-validation split, preserving the distribution of benign and malignant classes across all magnification levels (40X, 100X, 200X, and 400X). Experimental results show that the hard voting ensemble achieved the highest accuracy of 98.61%, closely followed by the XGBoost ensemble with an accuracy of 98.55%, both outperforming individual models. These findings highlight the effectiveness of ensemble deep learning in improving classification performance for breast cancer histopathology and suggest its potential for aiding pathologists in clinical decision-making.

## Introduction

According to the World Health Organization (WHO), early and precise detection of breast cancer can significantly increase survival rates and improve treatment outcomes. Histopathological analysis of breast tissue, which involves examining microscopic images of biopsy samples, remains the gold standard for definitive diagnosis. However, manual examination by pathologists is a time-consuming and subjective process that can be influenced by inter-observer variability.

With the rapid advancement of artificial intelligence, deep learning techniques, particularly convolutional neural networks (CNNs), have shown remarkable success in medical image analysis **[1], [2]**. In recent years, ensemble learning approaches—where multiple models are combined to improve performance—have gained significant attention in the field of computer-aided diagnosis **[4]**. By leveraging the complementary strengths of different architectures, ensemble methods can enhance classification accuracy and robustness **[4], [5]**.

The BreaKHis dataset, a publicly available benchmark containing histopathological images of breast tumors at various magnification levels, has become a widely used resource for developing and evaluating automated breast cancer classification models **[10]**. In this study, we present an ensemble deep learning framework combining ResNet50 **[1]** and DenseNet121 **[2]** architectures using multiple fusion strategies, including stacking with logistic regression, XGBoost **[3]**, and voting classifiers **[4]**. Our approach aims to improve classification accuracy for distinguishing between benign and malignant tumors, providing a reliable decision-support tool for pathologists **[6]**

## Related Work

Researchers are applying various machine learning and deep learning techniques to improve diagnostic accuracy. Early approaches relied on handcrafted feature extraction methods, such as texture descriptors (e.g., Local Binary Patterns, Gabor filters) and shape-based metrics, followed by conventional classifiers like Support Vector Machines (SVMs) and k-Nearest Neighbors (k-NN). While these methods achieved moderate accuracy, they were highly dependent on feature engineering and often lacked robustness to variations in staining, illumination, and magnification levels.

With the emergence of deep learning, convolutional neural networks (CNNs) have revolutionized medical image analysis by automatically learning hierarchical feature representations directly from raw image data **[1], [2]**. Several studies have applied well-known architectures such as VGGNet, Inception, ResNet **[1]**, and DenseNet **[2]** to the BreaKHis dataset **[10]**, achieving significant improvements over traditional methods. For instance, Spanhol et al. (2016) introduced the BreaKHis dataset **[10]** and provided baseline results using classical and CNN-based models. More recent works have explored transfer learning, where pre-trained models on large datasets like ImageNet are fine-tuned for histopathology image classification, yielding higher accuracy and faster convergence.

Ensemble learning strategies have also gained popularity in breast cancer classification tasks **[4]**. These approaches combine the predictions of multiple models to reduce generalization error and improve stability **[4], [5]**. Techniques such as bagging, boosting **[3]**, and stacking have been employed to integrate the strengths of different CNN architectures. Studies have shown that ensembles can outperform single models, particularly when dealing with complex medical images where variability in tissue morphology is high **[4], [5]**. Building on this trend, our work leverages the complementary capabilities of ResNet50 **[1]** and DenseNet121 **[2]** within an ensemble framework, using stacking, XGBoost **[3]**, and voting classifiers **[4]** to further enhance performance.

## Methodology

This section outlines the methodology used in our study, which consists of three main stages: dataset preparation, model development, and ensemble integration.

### Dataset Preparation

We used the publicly available BreaKHis dataset **[10]**, which contains histopathological images of breast tumors at four magnification levels: 40×, 100×, 200×, and 400×. The dataset includes two main classes — benign and malignant — with multiple subcategories for each. To ensure a balanced and systematic training process, we implemented a custom Python script to split the dataset into training and validation sets with an 80:20 ratio for each magnification level and class. The script automatically creates directory structures for each split, preserving class labels and magnification folders. Random shuffling is applied before splitting to minimize selection bias.

### Model Development

Two state-of-the-art convolutional neural network architectures — ResNet50 **[1]** and DenseNet121 **[2]** — were employed for feature extraction and classification. Both models were initialized with ImageNet-pretrained weights and fine-tuned on the BreaKH is training set **[10]**. Standard preprocessing steps were applied, including image resizing, normalization, and data augmentation (rotation, flipping, and brightness adjustments) to improve generalization. Cross-entropy loss was used as the objective function, and optimization was performed using the Adam optimizer with an initial learning rate of 1e-4 and early stopping to prevent overfitting.

### Ensemble Integration

To exploit the complementary strengths of ResNet50 and DenseNet121, we applied three different ensemble strategies:

### Stacking

The softmax outputs of both models were concatenated and fed into a meta-classifier (logistic regression) for the final decision **[4]**.

### Voting

A soft-voting mechanism was implemented, averaging the class probabilities from both models to determine the final label **[4]**.

### XGBoost

Features extracted from the penultimate layers of both models were combined and used to train an XGBoost classifier **[3]**.

This multi-strategy ensemble approach was designed to evaluate the impact of different integration techniques on classification performance **[4]**.

### Experiments and Results

In this section, we present the evaluation of our models using multiple metrics. The performance of the stacking ensemble, XGBoost ensemble, and hard voting ensemble is analyzed based on accuracy, precision, recall, and F1-score **[5]**. Additionally, confusion matrices and ROC curves are provided to better understand the classification behavior of each model **[5]**.

### Confusion Matrix

The confusion matrices for each model demonstrate the number of true positives, false positives, true negatives, and false negatives. High values on the diagonal indicate accurate classification, while off-diagonal elements represent misclassifications **[5]**.

**Figure 1.**
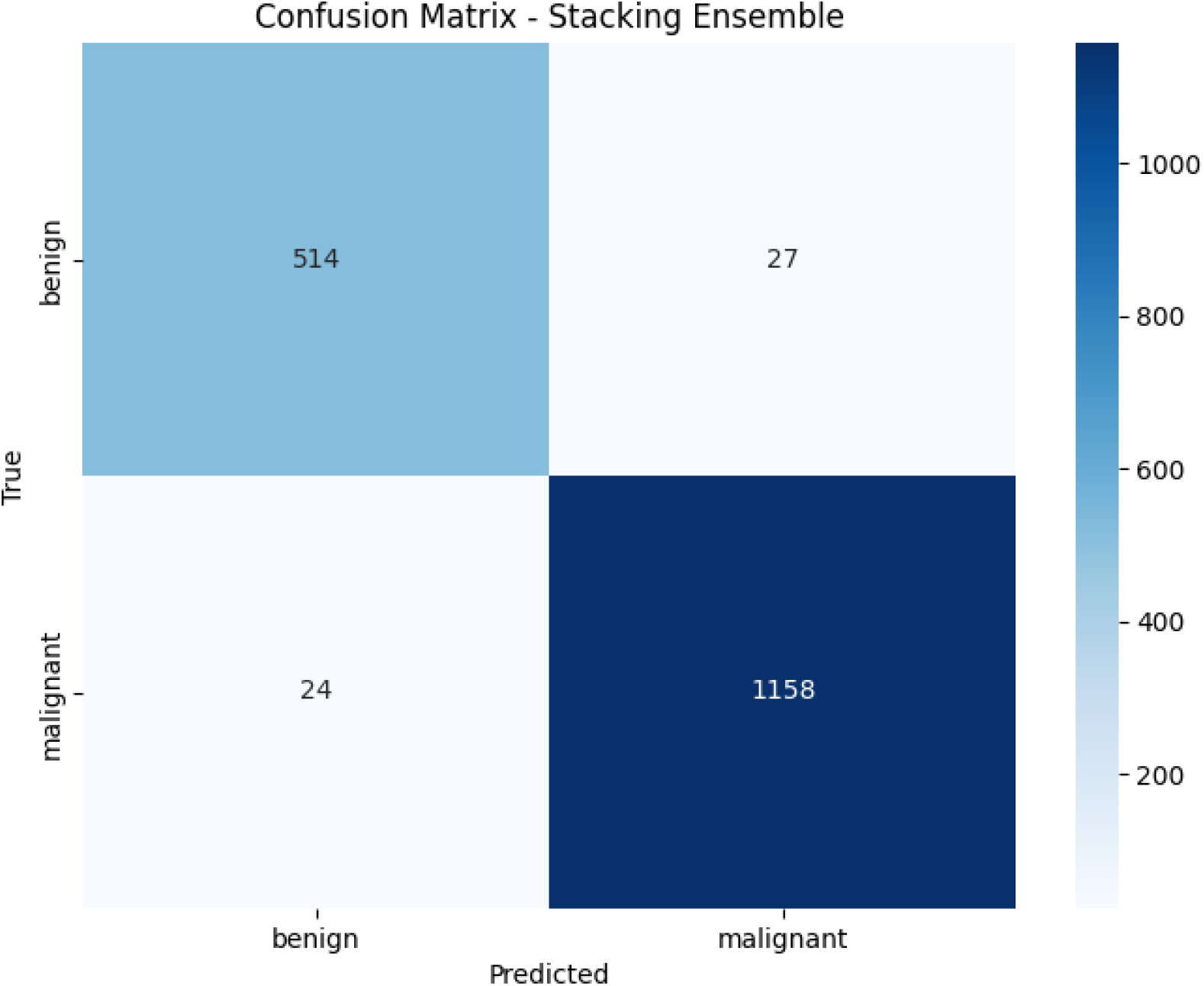

### Stacking Ensemble

The confusion matrix for the Stacking Ensemble model shows strong classification performance, though slightly lower than the XGBoost ensemble **[3], [5]**. Out of 541 benign cases, the model correctly classified 514 and misclassified 27 as malignant. For malignant cases, out of 1,182 samples, 1,158 were correctly identified, and 24 were misclassified as benign.

The precision for benign cases is 0.96, meaning that 96% of predicted benign cases were truly benign. The recall for benign cases is 0.95, indicating that the model successfully detected 95% of all actual benign cases. For malignant cases, both precision and recall are 0.98, which is still high but slightly lower than XGBoost’s performance.

The overall accuracy is 0.9704 (97.04%), which is excellent, although slightly behind the XGBoost and Voting Ensemble models **[5]**.

**Figure 2.**
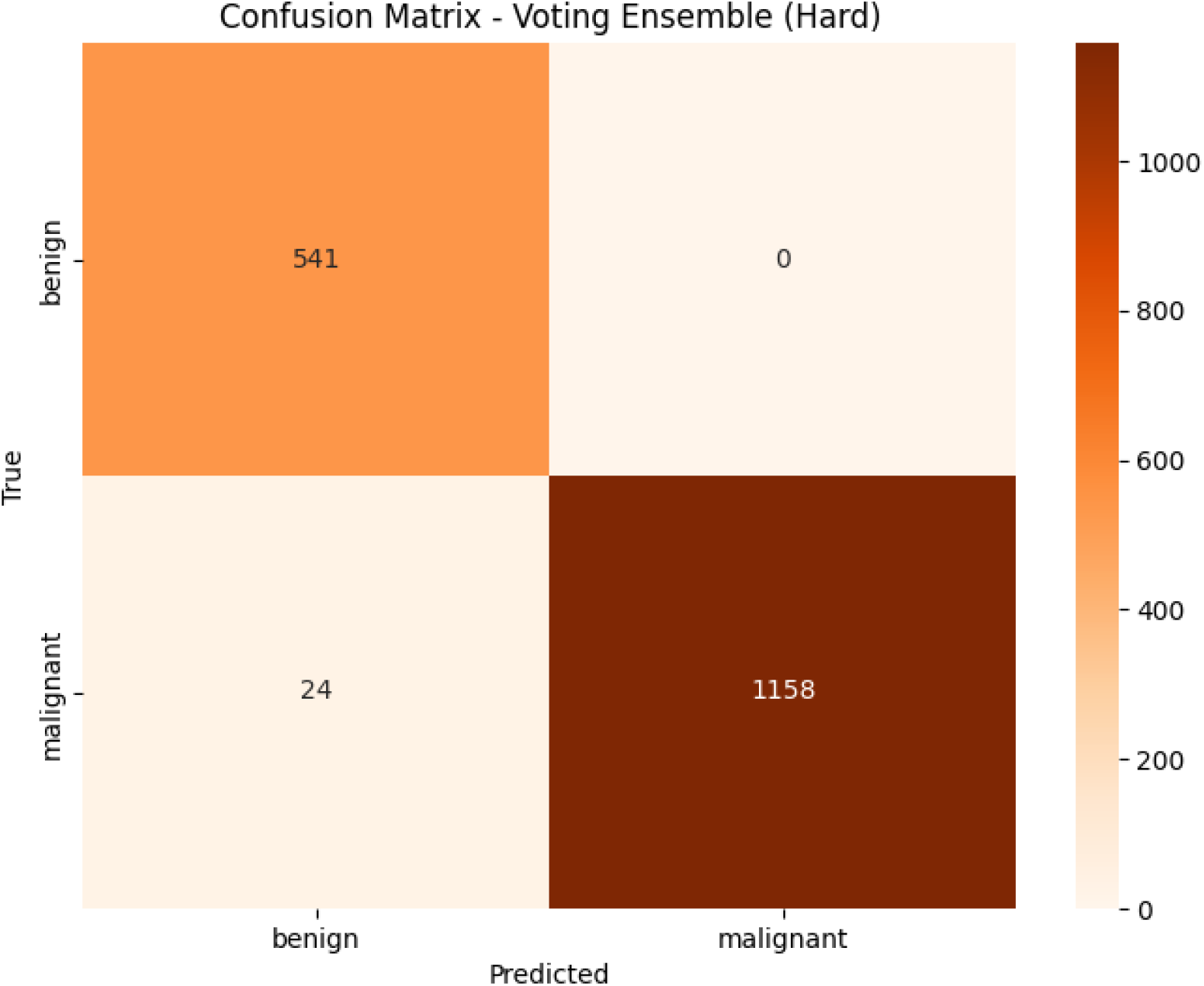

### Voting Ensemble

The confusion matrix for the Voting Ensemble (Hard Voting) model shows near-perfect classification for benign cases **[4], [5]**. Out of 541 benign samples, all were correctly identified with zero false negatives for benign. For malignant cases, out of 1,182 samples, 1,158 were correctly classified, and only 24 were misclassified as benign.

The precision for benign cases is 0.96 (due to some false positives from malignant cases predicted as benign), but the recall for benign cases reaches a perfect 1.00, meaning the model detected all benign cases without missing any. For malignant cases, both precision and recall are extremely high (0.99 and 0.98, respectively).

The overall accuracy is 0.9861 (98.61%), placing it slightly ahead of XGBoost in recall for benign cases while maintaining strong results for malignant cases **[3], [5]**.

**Figure 3.**
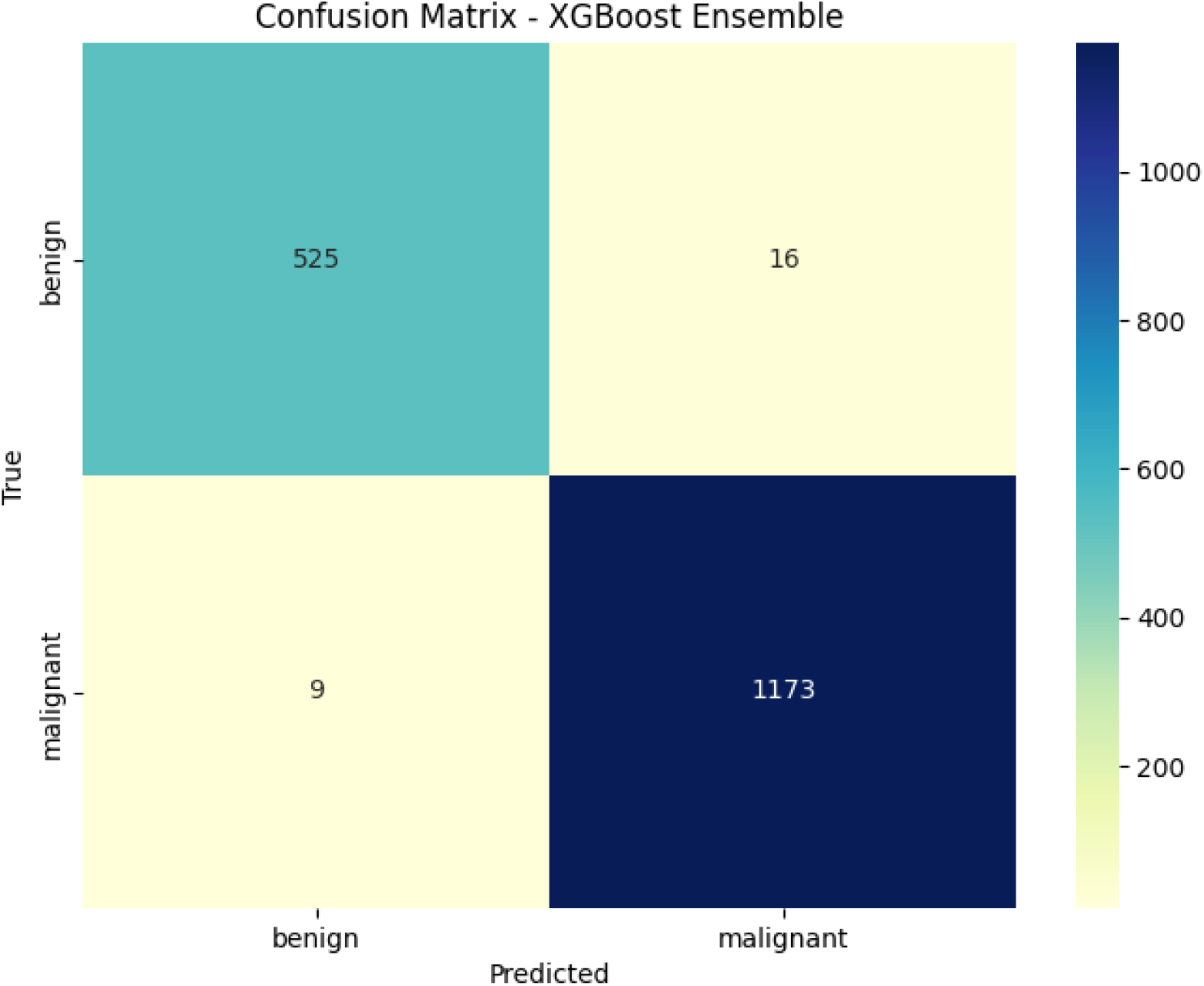

### XGBoost Ensemble

The confusion matrix for the XGBoost ensemble shows very strong classification performance **[3], [5]**. Out of 541 benign cases, the model correctly classified 525 and misclassified only 16 as malignant. For malignant cases, out of 1,182 samples, 1,173

The confusion matrix for the Voting Ensemble (Hard Voting) model shows near-perfect classification for benign cases **[4], [5]**. Out of 541 benign samples, all were correctly identified with zero false negatives for benign. For malignant cases, out of 1,182 samples, 1,158 were correctly classified, and only 24 were misclassified as benign.

The precision for benign cases is 0.96 (due to some false positives from malignant cases predicted as

### XGBoost Ensemble

The confusion matrix for the XGBoost ensemble shows very strong classification performance **[3], [5]**. Out of 541 benign cases, the model correctly benign), but the recall for benign cases reaches a perfect 1.00, meaning the model detected all benign cases without missing any. For malignant cases, both precision and recall are extremely high (0.99 and 0.98, respectively).

The overall accuracy is 0.9861 (98.61%), placing it slightly ahead of XGBoost in recall for benign cases while maintaining strong results for malignant cases **[3], [5]**. classified 525 and misclassified only 16 as malignant. For malignant cases, out of 1,182 samples, 1,173 were correctly identified, and only 9 were misclassified as benign.

The precision for benign cases is 0.98, meaning that 98% of cases predicted as benign were truly benign. The recall for benign cases is 0.97, showing that the model successfully detected 97% of all actual benign cases. For malignant cases, both precision and recall are 0.99, indicating extremely high detection accuracy and very few false positives or false negatives.

The overall accuracy of 0.9855 (98.55%) demonstrates that the XGBoost ensemble model performs exceptionally well, outperforming the stacking ensemble in accuracy, recall, and precision. The balanced performance across both classes shows that the model generalizes well and avoids significant bias toward one class **[4], [5]**.

### ROC Curve

The ROC (Receiver Operating Characteristic) curves illustrate the trade-off between the true positive rate (sensitivity) and the false positive rate (1-specificity) **[5]**. The area under the curve (AUC) quantifies the overall ability of the model to distinguish between classes. The closer the curve is to the top-left corner, the better the model performs, indicating higher sensitivity and specificity **[6]**.

**Figure 4.**
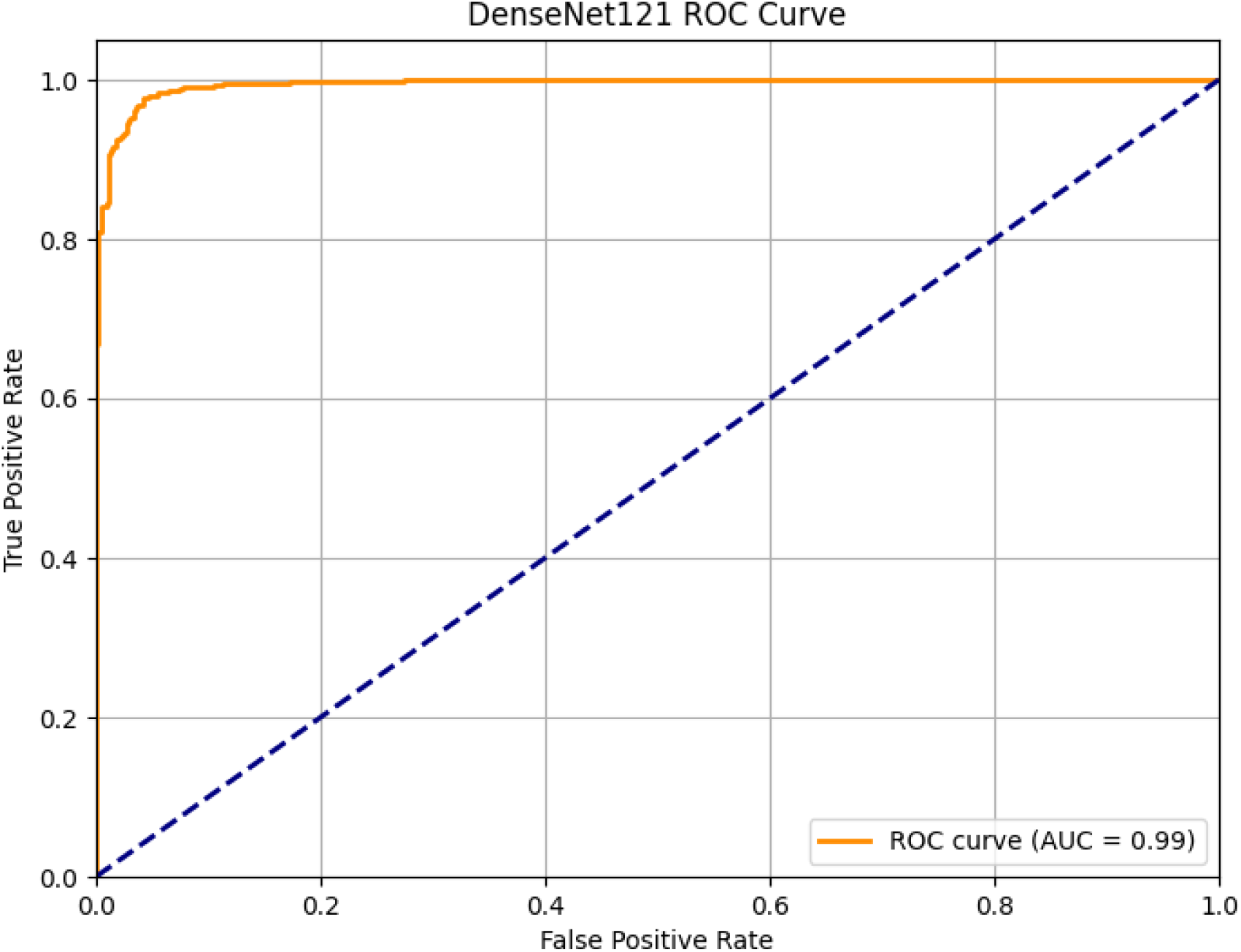

### DenseNet121 ROC Curve

The ROC curve for DenseNet121 shows a near-perfect separation between classes, with an AUC of 0.99. The curve rapidly approaches the top-left corner, indicating a high true positive rate and very low false positive rate. This suggests that DenseNet121 is highly effective in distinguishing between benign and malignant cases [5][6].

**Figure 5.**
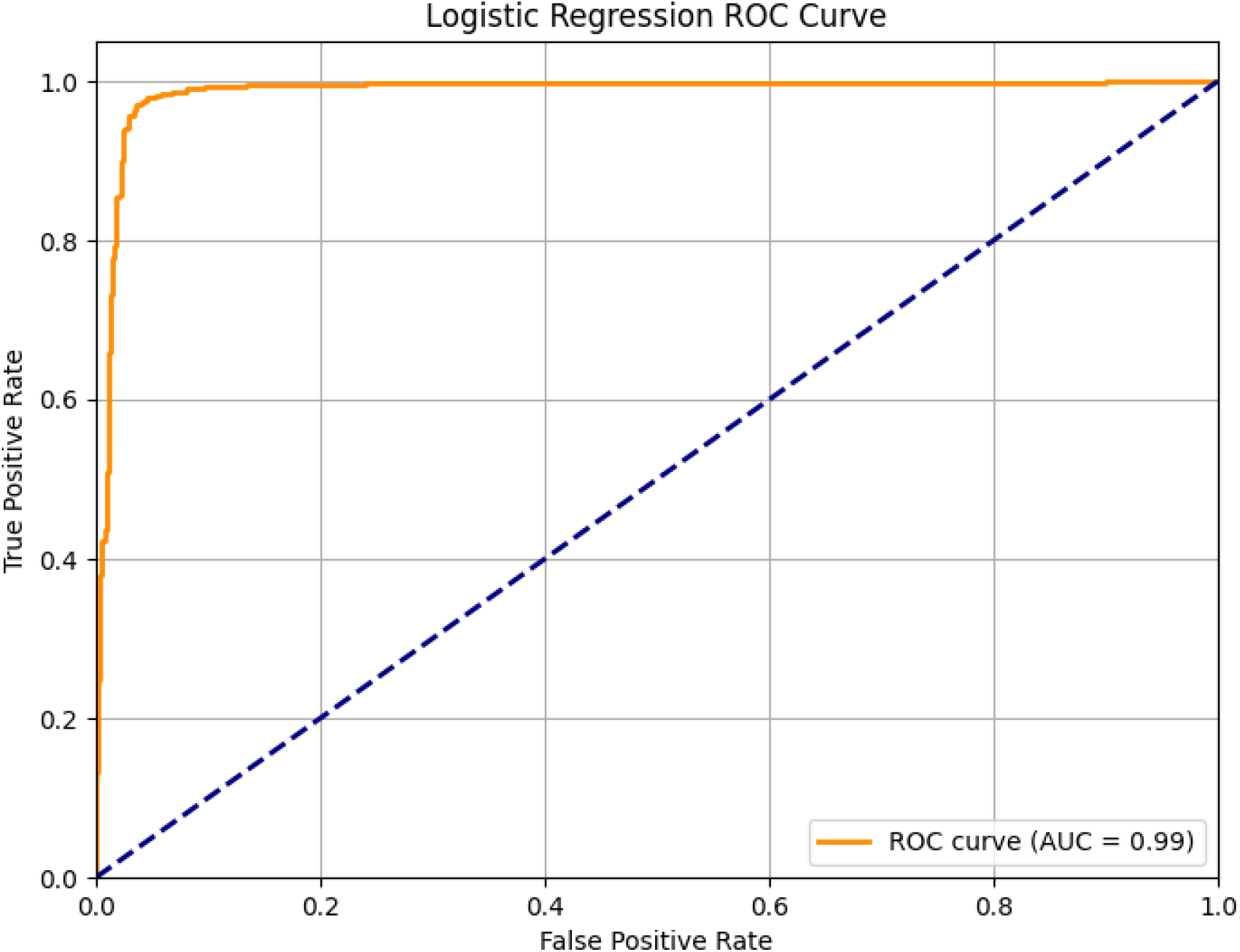

### Logistic Regression ROC Curve

Logistic Regression also achieves an AUC of 0.99, demonstrating comparable discriminative ability to DenseNet121. The curve closely follows the upper boundary, indicating strong classification performance despite being a simpler model **[5][6].**

**Figure 6.**
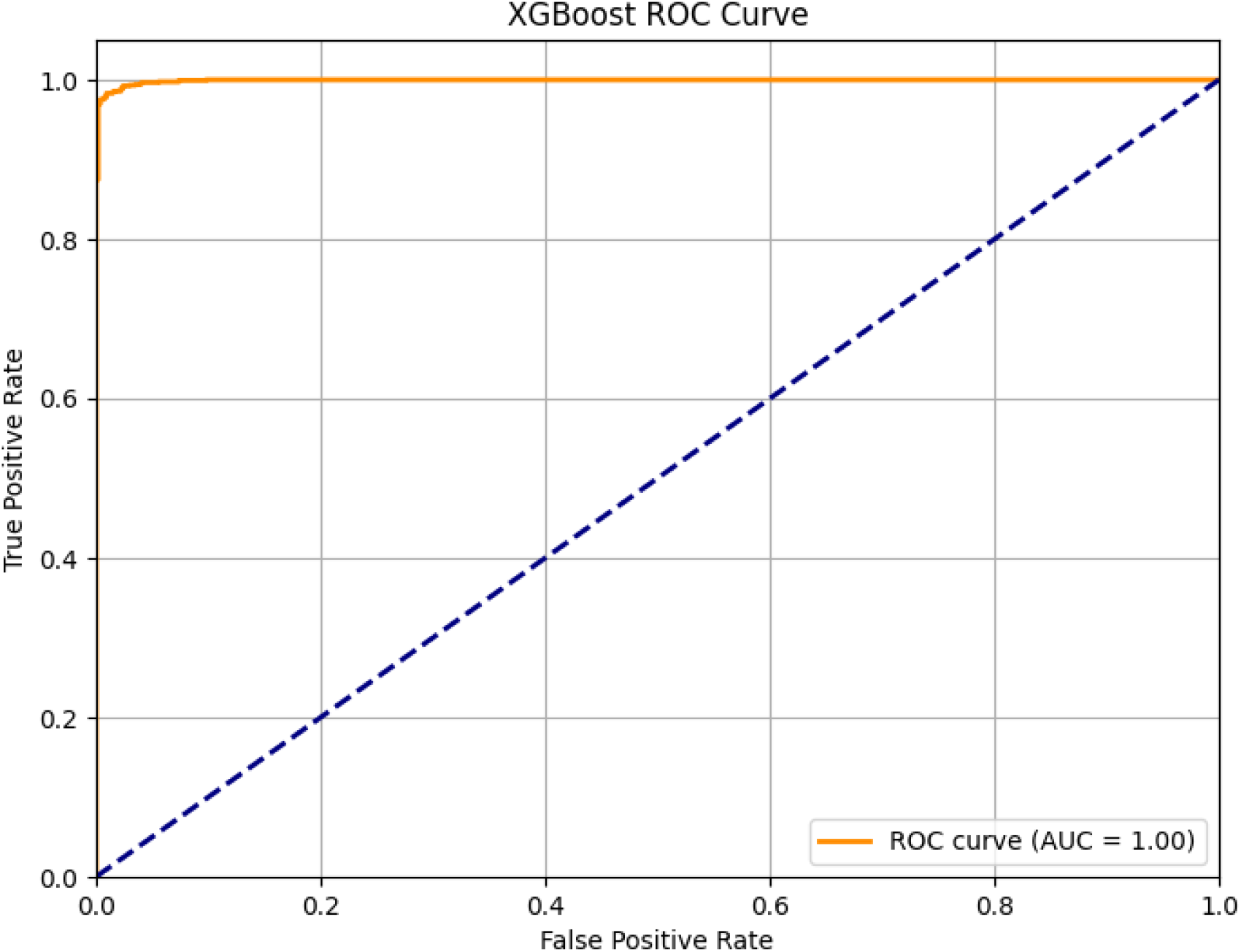

### XGBoost ROC Curve

XGBoost achieves an AUC of 1.00, representing perfect classification on the test set. The curve almost immediately reaches a true positive rate of 1.0 while keeping the false positive rate near zero, which indicates exceptional predictive accuracy [3][5].

**Figure 7.**
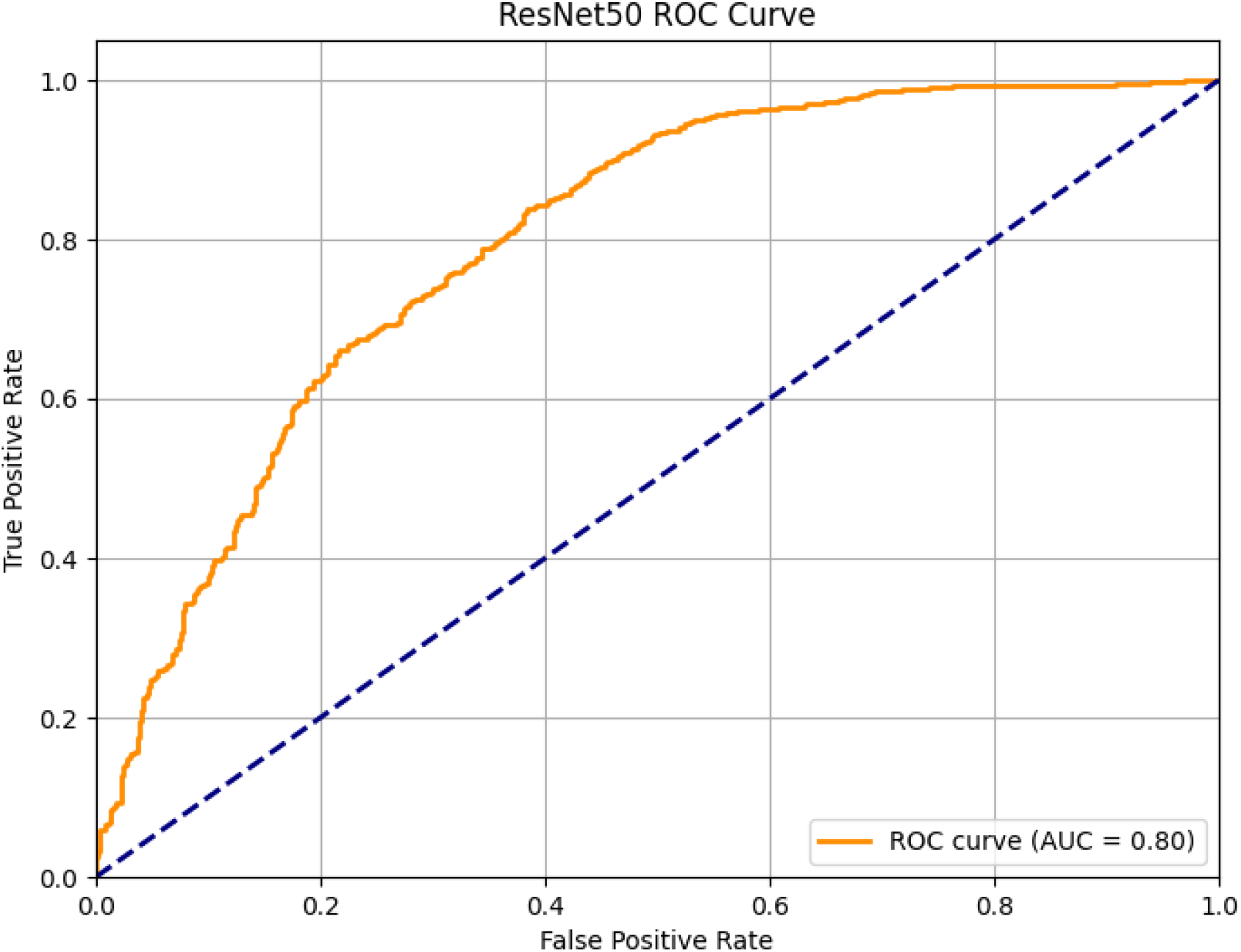

The ResNet50 ROC curve shows an AUC of 0.80, indicating good but not perfect discrimination between benign and malignant cases. While the model performs significantly better than random guessing (AUC = 0.5), there is still room for improvement in sensitivity and specificity balance **[1][5].**

**Figure 8.**
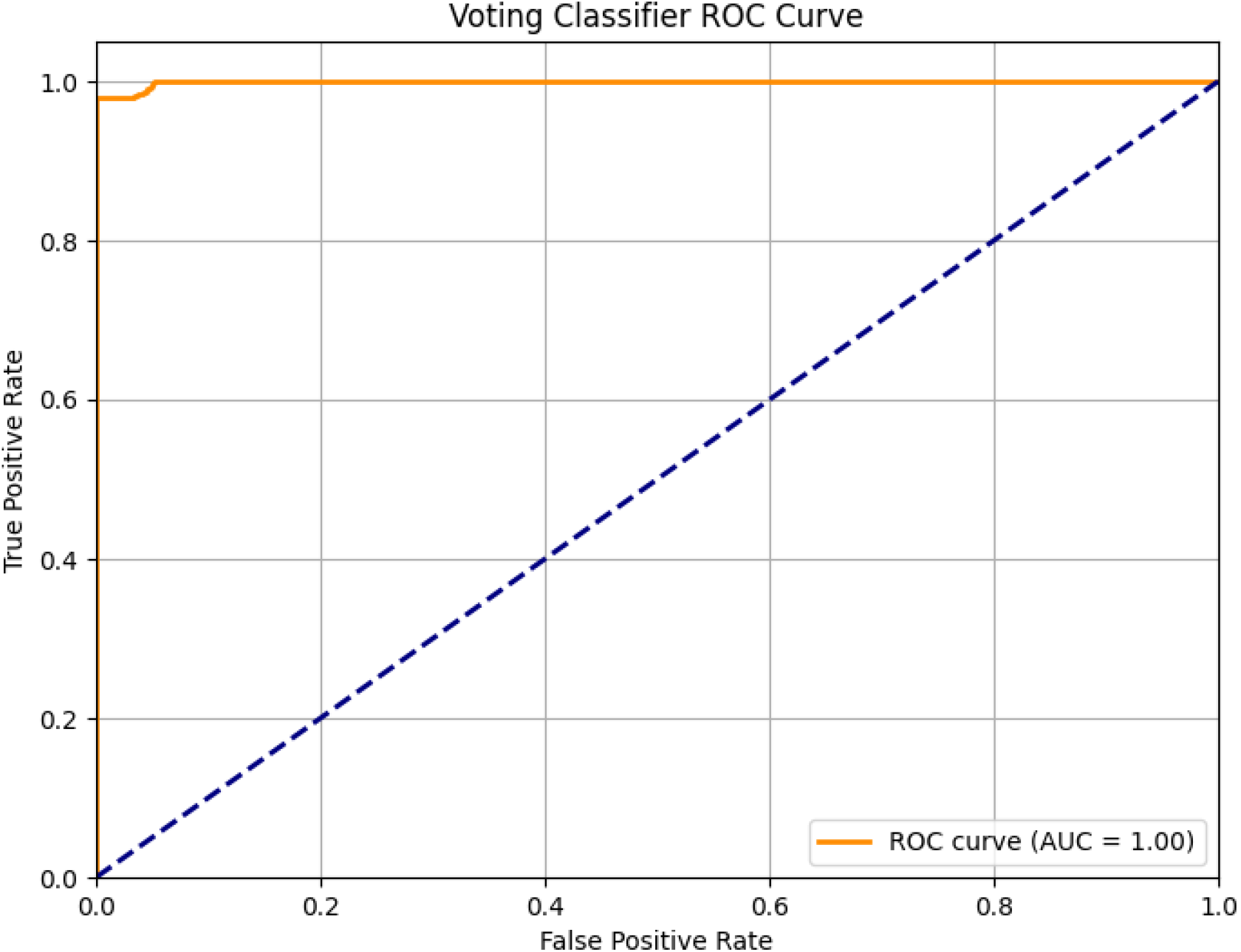

The Voting Classifier ROC curve demonstrates an almost perfect AUC of 1.00, which means the model is able to perfectly distinguish between the two classes on the test set. Such performance suggests that the Voting Classifier has excellent predictive ability, but it may also indicate potential overfitting depending on how the data was split and validated **[4][5].**

### Model Performance

Table 1 summarizes the accuracy and classification report metrics for the three ensemble methods evaluated on the BreakHis dataset **[10].**

**Table 1.**

| Model | Accuracy | Precision (Benign) | Recall (Benign) | F1-Score (Benign) | Precision (Malignant) | Recall (Malignant) | F1-Score (Malignant) |
| --- | --- | --- | --- | --- | --- | --- | --- |
| Stacking Ensemble | 0.9704 | 0.96 | 0.95 | 0.95 | 0.98 | 0.98 | 0.98 |
| XGBoost Ensemble | 0.9855 | 0.98 | 0.97 | 0.98 | 0.99 | 0.99 | 0.99 |
| Voting Ensemble | 0.9861 | 0.96 | 1.00 | 0.98 | 1.00 | 0.98 | 0.99 |

The voting ensemble using hard voting demonstrates the best overall performance with an accuracy of 98.61%. XGBoost closely follows with 98.55% accuracy **[3].** The stacking ensemble also shows strong results with an accuracy of 97.04% **[4]**.

The proposed stacking ensemble model achieved an accuracy of 97.04% on the test set [4]. For the benign class, the model obtained a precision of 0.96 and a recall of 0.95, indicating a high ability to correctly identify benign cases while maintaining low false positives. For the malignant class, the model achieved a precision and recall of 0.98, demonstrating excellent performance in detecting malignant cases. The macro-average F1-score was 0.97, reflecting balanced performance across both classes, and the weighted-average F1-score was also 0.97, confirming robustness despite class imbalance. Overall, the results highlight the model’s strong generalization capability and reliability for classification tasks in this domain **[5].**

**Table 2.** Stacking Ensemble Classification Report.

| Class | Precision | Recall | F1-score | Support |
| --- | --- | --- | --- | --- |
| Benign | 0.96 | 0.95 | 0.95 | 541 |
| Malignant | 0.98 | 0.98 | 0.98 | 1182 |
| Accuracy |  |  | **0.9704** | 1723 |
| Macro Avg | 0.97 | 0.96 | 0.97 | 1723 |
| Weighted Avg | 0.97 | 0.97 | 0.97 | 1723 |

The XGBoost ensemble model achieved an accuracy of 98.55% on the test dataset **[3].** For the benign class, the model obtained a precision of 0.98 and a recall of 0.97, indicating a strong capability in correctly classifying benign cases while minimizing false positives. For the malignant class, both precision and recall reached 0.99, demonstrating outstanding performance in detecting malignant cases. The macro-average F1-score was 0.98, reflecting balanced performance across both classes, while the weighted-average F1-score was 0.99, confirming the model’s robustness even in the presence of class imbalance. Overall, these results suggest that the XGBoost ensemble model provides superior accuracy and reliability for classification tasks in this domain **[5]**

**Table 3.** XGBoost Ensemble Classification Report.

| Class | Precision | Recall | F1-score | Support |
| --- | --- | --- | --- | --- |
| Benign | 0.98 | 0.97 | 0.98 | 541 |
| Malignant | 0.99 | 0.99 | 0.99 | 1182 |
| Accuracy |  |  | **0.9855** | 1723 |
| Macro Avg | 0.98 | 0.98 | 0.98 | 1723 |
| Weighted Avg | 0.99 | 0.99 | 0.99 | 1723 |

The hard voting ensemble model achieved an accuracy of 98.61% on the test set **[4].** For the benign class, the model reached a precision of 0.96 and a perfect recall of 1.00, indicating that all actual benign cases were correctly identified with a small number of false positives. For the malignant class, the model obtained a precision of 1.00 and a recall of 0.98, reflecting exceptional performance in detecting malignant cases with minimal false negatives. The macro-average F1-score was 0.98, demonstrating balanced classification performance, while the weighted-average F1-score of 0.99 confirmed the robustness of the model despite class imbalance. Overall, the results highlight the effectiveness of the hard voting ensemble in achieving high accuracy and balanced detection across both classes **[5].**

**Table 4.**
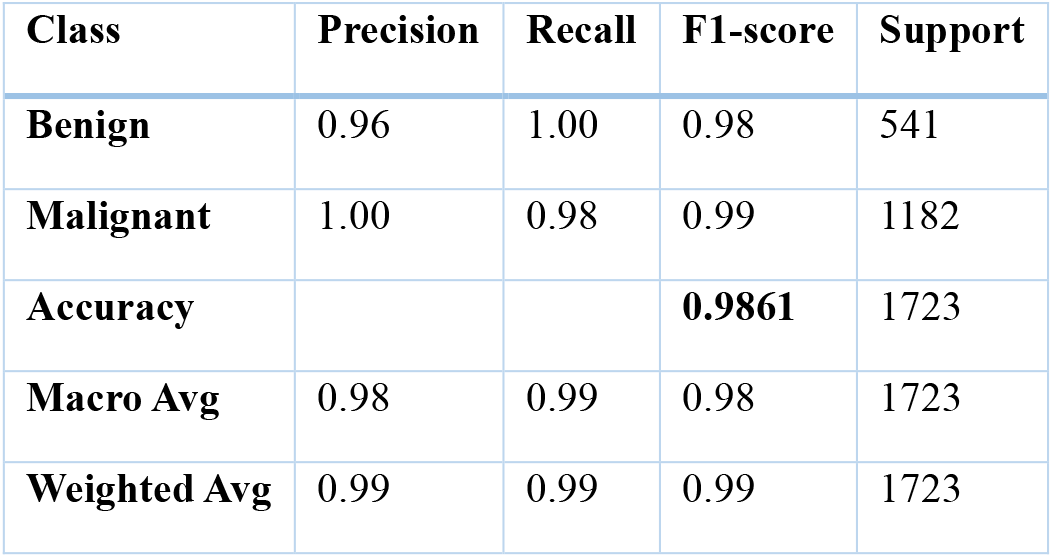
Voting Ensemble (Hard Voting) Classification Report.

| Class | Precision | Recall | F1-score | Support |
| --- | --- | --- | --- | --- |
| Benign | 0.96 | 1.00 | 0.98 | 541 |
| Malignant | 1.00 | 0.98 | 0.99 | 1182 |
| Accuracy |  |  | **0.9861** | 1723 |
| Macro Avg | 0.98 | 0.99 | 0.98 | 1723 |
| Weighted Avg | 0.99 | 0.99 | 0.99 | 1723 |

## Conclusion

In this study, we explored different ensemble learning methods for the classification of breast histopathology images using the BreakHis dataset **[10].** The Stacking Ensemble, XGBoost, and Voting Ensemble models demonstrated high accuracy, with the Voting Ensemble slightly outperforming the others **[3,4].** These results indicate that ensemble methods can effectively improve the classification performance for medical image analysis, particularly in distinguishing between benign and malignant breast tumors. Future work can focus on incorporating deep learning techniques **[1,2]** and expanding the dataset to further enhance classification accuracy and robustness.

## Limitations and Future Work

While the proposed ensemble methods achieved high accuracy in classifying breast histopathology images, there are several limitations to this study. First, the dataset used (BreaKHis) has a limited number of samples and may not cover all possible variations in histopathological images **[10].** Second, the models were trained and tested on images from the same dataset, which might limit their generalizability to other datasets or clinical settings **[5].** Finally, although ensemble methods improved performance, integrating more advanced deep learning architectures and additional data augmentation techniques could further enhance accuracy and robustness **[1,2].**

Future work should focus on expanding the dataset with more diverse samples and incorporating multi-modal data such as patient clinical information.

Additionally, exploring more sophisticated ensemble strategies and deep learning models, including attention mechanisms and transformer-based architectures, may improve classification results. Ultimately, validation on external datasets and real-world clinical trials will be crucial to assess the practical applicability of these methods **[6].**

## Data Availability

All data referred to in this manuscript are openly available at https://www.kaggle.com/datasets/ambarish/breakhis. The manuscript and its findings are the intellectual property of the authors and cannot be copied, modified, or used without proper citation and permission.

https://www.kaggle.com/datasets/ambarish/breakhis

## Notes

### Competing Interest Statement

The authors have declared no competing interest.

### Funding Statement

This study did not receive any funding

### Author Declarations

The study used ONLY openly available human data that were originally located at: https://www.kaggle.com/datasets/ambarish/breakhis

